# Development and pilot evaluation of ACE-T, a nurse-focused delirium initial-response tool, across three health systems

**DOI:** 10.64898/2026.11.08.26367532

**Authors:** Alexandra Cours, Johannes Trabert, Maggie Higgins, Umar Saleem, Elizabeth Sampson, Nannette Storr-Street, Alasdair MacLullich

**Affiliations:** Section of Geriatric Medicine, Primary Care and Population Health, Department of Medicine, Stanford University School of Medicine, Palo Alto, CA, USA; Medizinisch-Geriatrische Klinik, AGAPLESION Markus Krankenhaus, Frankfurt am Main, Germany; Medicine of the Elderly, Royal Infirmary of Edinburgh, NHS Lothian, Edinburgh, United Kingdom; Academic Centre for Healthy Ageing, Centre for Psychiatry and Mental Health, Wolfson Institute of Population Health, Queen Mary University of London, London, United Kingdom; Ageing and Health, Usher Institute, University of Edinburgh, Edinburgh, United Kingdom; Inpatient Geriatrics, Stanford Health Care, Palo Alto, CA, USA

## Abstract

**Background:** A positive delirium screen should trigger timely assessment and care, but the initial bedside response is not always clearly defined. ACE-T (Acute Triggers / Patient Experience / Treatments) is a nurse-led tool (www.theace-t.com) that sets out initial care actions and supports documentation and team communication.

**Objectives:** To assess perceived usability and utility, refine the tool, and describe documentation before and after its introduction at three hospitals.

**Methods:** In this developmental pilot, 53 multidisciplinary staff completed a structured survey and provided free-text feedback. We also reviewed a non-random sample of 60 records: 10 before and 10 after ACE-T introduction at each site. Prespecified delirium-care actions documented within 48 hours of the first positive delirium screen or documented diagnosis were summarised descriptively by site.

**Results:** ACE-T was rated easy or very easy to use by 75% (39/52), clear or very clear by 77% (41/53), and very or extremely helpful for improving delirium care by 71% (37/52). Feedback guided revisions to clarify professional roles, fit clinical workflows and strengthen person-centred prompts. Documentation of delirium or suspected delirium was more frequent after introduction at all three sites: 1/10 before versus 10/10 after in Germany, 0/10 versus 10/10 in the UK, and 3/10 versus 9/10 in the USA. Changes in documentation of other assessments varied by site.

**Conclusion:** ACE-T is a promising nurse-led tool for initial delirium care. Positive staff ratings and more frequent delirium documentation support its further development and prospective evaluation. It offers nurses a practical starting point for what to do next when delirium is detected.

## Introduction

Delirium care involves three core elements: detection, treatment, and prevention.^1-4^ Standard approaches for detection and prevention are well established in the literature.^5-8^ By contrast, delirium treatment is much less well defined.^1,9^ Across guidelines and other sources, approaches to delirium treatment commonly describe a package of multiple actions including, for example, identifying and addressing triggers, managing symptoms, and promoting recovery.^2,3,10-13^ Yet these approaches show substantial variations in content and operationalization.

In guidelines, delirium treatment has a range of labels, including ‘multicomponent non-pharmacological interventions’, a ‘multicomponent program’, ‘non-drug strategies’, and ‘established pathways of good care’. The content ranges from two brief recommendations to a 15-component program.^2,3,10-13^ Most guidelines do not define which professional groups are responsible for what actions. Exceptions are the American Psychiatric Association 2025 guideline,^12^ which notes that nurses “deliver or assure delivery” of most non-pharmacological interventions, and the German guideline^13^ which places the acute etiological work-up at specialist-physician level and allocates non-pharmacological interventions to nursing, occupational therapy and physiotherapy by problem domain. Even these exceptions are partial: neither defines the sequence or timing of actions required of nurses in the hours immediately after a positive screen.

Outside guidelines, delirium treatment is described in research studies, book chapters, order sets, and clinical pathways.^14-16^ As with guidelines, these sources vary considerably in sequence, timing, role of different disciplines (which is often not specified), and documentation. A consequence of this lack of standardization is that clinical teams vary substantially in how they respond after a positive delirium screen. Studies show variation and deficiencies in identifying modifiable triggers, assessing and managing patient distress, and communicating the diagnosis to patients and families.^17-21^

One potentially useful term for the package of actions involved in treatment is ‘multidomain’ delirium treatment.^1^ The term avoids the misleading division of clinical delirium treatment into distinct ‘non-pharmacological’ and ‘pharmacological’ elements. This is important because guidelines advise against giving psychotropic drugs routinely, but they do permit brief, targeted use if necessary for intractable distress or safety considerations.^2,3,10-13^ This means that the use of drugs is simply one optional tool in a wider set of possible actions, not an implied treatment category of its own. ‘Multidomain’ also separates this package-based approach from single-intervention strategies, such as drug-based or music-based interventions. Here we use the term multidomain to describe mainstream delirium treatment as used in routine clinical care.

Although the roles of the healthcare team overlap, it is reasonable for different disciplines to focus on the aspects of delirium treatment best suited to each, as stated in the American and German guidelines, and in some published studies.^12,13,22-26^ In many healthcare systems, nurses are the main group carrying out delirium screening, meaning that they are in the ideal position to respond as soon as a screen is positive. Nurses also have a central role in the ongoing care of patients with delirium: managing interventions, monitoring distress, communicating with families, and keeping patients safe. Given the urgency of delirium treatment and the central role of nurses in detection and care, a practical nurse-focused tool is needed to guide the first steps after a positive screen. Such a tool could help standardize these first steps, alongside the complementary medical and other roles in delirium care. Additionally, several studies have identified gaps in escalation and clinical action following a positive delirium screen,^27-30^ and nurses are well placed to take rapid action and help initiate involvement of other team members.

Here we describe the development and preliminary piloting of the ACE-T tool (ACute Triggers / patient Experience / Treatments). This is a brief nurse-led multidomain delirium treatment tool intended to standardize initial bedside nursing actions following a positive delirium screen. Our aims were: (1) to assess the usability and perceived utility of the tool among multidisciplinary clinical staff; (2) to describe how nurse-led delirium-care actions were documented after a positive screen; and (3) to identify implementation and measurement lessons for a future, more definitive evaluation.

## Methods

### Study design and setting

We used a staged, multicenter developmental pilot design comprising development of a provisional ACE-T tool; a pre-introduction staff survey and revision; and site-led pilot use with a descriptive before-and-after review of clinical documentation. The work took place at Stanford Hospital (Stanford, USA), AGAPLESION Markus Krankenhaus (Frankfurt, Germany), and the Royal Infirmary of Edinburgh (Edinburgh, UK). Baseline record-review data were collected in summer 2025 and post-introduction data in autumn and winter 2025-2026. The study assessed staff perceptions, documentation patterns, and measurement issues; it was not designed to estimate clinical effectiveness.

### Development of the provisional ACE-T tool

AM, MH, AC and JT iteratively drafted the provisional tool drawing from published delirium guidelines and clinical care standards,^2,3,11^ the structure and content of the multidisciplinary Triggers, Investigations, Management and Engagement (TIME) bundle,^2^ and our clinical experience.

The ACE-T (www.theace-t.com) was developed to support nurse-led action in the first hours after a positive delirium screen or recognition of suspected delirium. The nursing focus was informed by evidence of deficiencies in post-screen clinical responses and by nurses’ key role in the early assessment, escalation and ongoing care of patients with delirium.^18,20,27,29,31,32^ The ACE-T organizes actions into the three domains of (1) ACute triggers, (2) patient Experience, and (3) Treatment and communication. These domains were chosen to prioritize early nursing actions while keeping the tool pragmatic.

The ACE-T prompts consideration of observations, review of whether clinically indicated investigations and medication review have occurred, assessment of distress (with the option of using a brief 4-point tool called the Quick Distress Assessment Tool (QDAT, Table 1))^21^ and the care environment, documentation of delirium, and communication with the clinical team and family. It does not require nurses to diagnose the cause or order investigations outside local policy, and it does not replace wider multidisciplinary care; the communication domain supports team involvement.

**Table 1.**
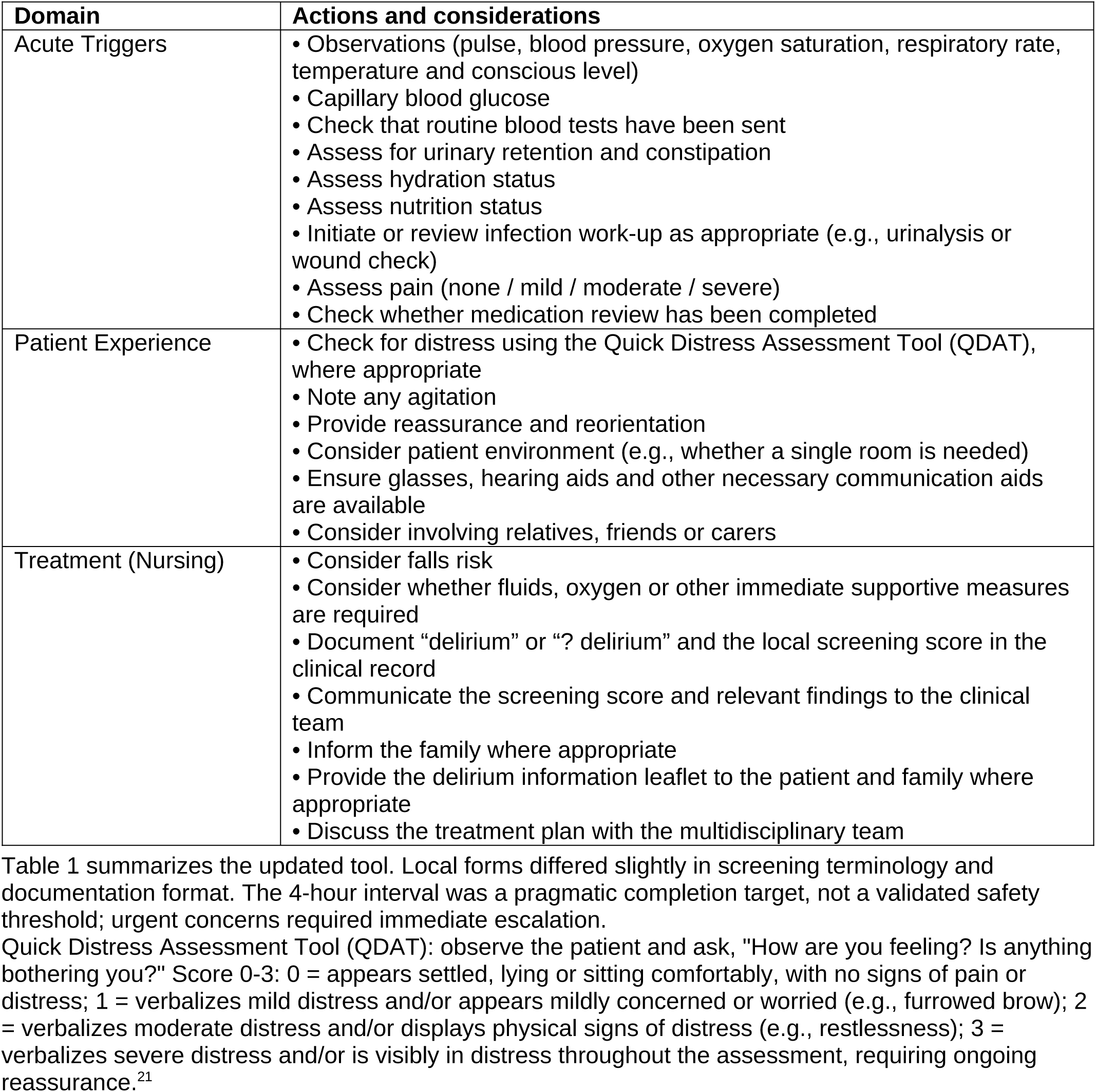
The ACE-T tool: Acute Triggers / Patient Experience / Treatments.

| **Domain** | **Actions and considerations** |
|---|---|
| Acute Triggers | • Observations (pulse, blood pressure, oxygen saturation, respiratory rate, temperature and conscious level)<br>• Capillary blood glucose<br>• Check that routine blood tests have been sent<br>• Assess for urinary retention and constipation<br>• Assess hydration status<br>• Assess nutrition status<br>• Initiate or review infection work-up as appropriate (e.g., urinalysis or wound check)<br>• Assess pain (none / mild / moderate / severe)<br>• Check whether medication review has been completed |
| Patient Experience | • Check for distress using the Quick Distress Assessment Tool (QDAT), where appropriate<br>• Note any agitation<br>• Provide reassurance and reorientation<br>• Consider patient environment (e.g., whether a single room is needed)<br>• Ensure glasses, hearing aids and other necessary communication aids are available<br>• Consider involving relatives, friends or carers |
| Treatment (Nursing) | • Consider falls risk<br>• Consider whether fluids, oxygen or other immediate supportive measures are required<br>• Document “delirium” or “? delirium” and the local screening score in the clinical record<br>• Communicate the screening score and relevant findings to the clinical team<br>• Inform the family where appropriate<br>• Provide the delirium information leaflet to the patient and family where appropriate<br>• Discuss the treatment plan with the multidisciplinary team |

Table 1 summarizes the updated tool. Local forms differed slightly in screening terminology and documentation format. The 4-hour interval was a pragmatic completion target, not a validated safety threshold; urgent concerns required immediate escalation.
Quick Distress Assessment Tool (QDAT): observe the patient and ask, "How are you feeling? Is anything bothering you?" Score 0-3: 0 = appears settled, lying or sitting comfortably, with no signs of pain or distress; 1 = verbalizes mild distress and/or appears mildly concerned or worried (e.g., furrowed brow); 2 = verbalizes moderate distress and/or displays physical signs of distress (e.g., restlessness); 3 = verbalizes severe distress and/or is visibly in distress throughout the assessment, requiring ongoing reassurance.<sup>21</sup>

Early drafts underwent revisions following feedback from nurses and physician colleagues to refine layout and content.

### Staff survey and revision

Before clinical use, local investigators invited a convenience sample of nurses, physicians, physician associates or assistants, and nurse practitioners to review the provisional tool and complete an anonymous paper questionnaire (Supplement 1). It recorded professional group and delirium-care experience. Four five-level items assessed ease of understanding and use, clarity, and perceived helpfulness for guiding and improving care. A yes/no item with free text asked whether any section was unclear or difficult, and an open question invited suggested changes. Ratings reflected perceived rather than observed usability. Participation was voluntary, and return indicated consent.

The project group reviewed the ratings and comments and revised the content and layout. The revised version retained the three domains, clarified nursing actions and medical escalation, simplified Acute Triggers, and expanded prompts about distress, reorientation, sensory aids, carers, and safety. Following feedback concerning various timescales for completion, it set a target of 4 hours from the positive screen for completing, initiating, or escalating applicable actions; urgent concerns required immediate action. The questionnaire is provided in Supplement 1, and the revised pilot version is shown in Table 1.

### Pilot introduction and record sampling

Each site introduced ACE-T through existing clinical teams without a shared training package or standardized procedure (Table 2). At Stanford, the geriatric Clinical Nurse Specialist introduced the paper tool on the Acute Care for Elders unit after a positive Confusion Assessment Method screen. Edinburgh used a paper tool in geriatric medicine wards. In Frankfurt, the medical ward delirium team used ACE-T after a positive 4AT and recorded actions electronically. We did not assess uptake, fidelity, or completion within 4 hours.

**Table 2.** Implementation and sampling context by site.

| **Site** | **Implementation and sampling features** | **Key interpretive caveat** |
|---|---|---|
| Frankfurt | Positive 4AT screen; tool introduced by the local investigator to the delirium team responsible for medical wards; documentation entered in existing delirium-team electronic records. | Baseline routine clinical items were abstracted from delirium-team records rather than comprehensive ward records. Apparent absence of vital signs, blood tests or hydration assessment should not be interpreted as absence of routine ward care. |
| Edinburgh | Tool introduced through local geriatric medicine nursing links on participating wards at the Royal Infirmary of Edinburgh; completed paper tools collected locally. Post-implementation sample came from geriatric units. | Ward mix changed between phases, limiting before-after interpretation. Post-implementation documentation may reflect geriatric-unit practice as well as tool use. |
| Stanford | Geriatric Clinical Nurse Specialist introduced paper tool on the Stanford Acute Care for Elders unit for patients with a positive CAM screen; completed forms were shared securely with the geriatric CNS or ACE-unit medical director. | Post-implementation sample came from a geriatric ACE unit; nursing scope and comfort with documenting "delirium" versus "? delirium" may differ from other sites. |

Abbreviations: 4AT, 4 A's Test; ACE, Acute Care for Elders; CAM, Confusion Assessment Method; CNS, Clinical Nurse Specialist.

Local reviewers identified 10 records before and 10 after introduction at each site (60 records). Eligible records contained a positive delirium screen or documented clinical diagnosis; screening instruments followed routine local practice. Case identification was not standardized or random, and samples were not intended to represent all eligible admissions. Post-introduction samples in Edinburgh and Stanford came from geriatric units, whereas baseline samples included more general medical wards.

### Record review

The index event was the earliest positive qualifying screen or, if none was recorded, the earliest explicit clinical documentation of delirium. A structured extraction form (Supplement 2) recorded age, sex, ward, time from admission to the index event, and documentation of prespecified care actions during the next 48 hours. These covered distress, pain, physiological contributors, delirium documentation, communication, written information, and patient-centered actions. Completed ACE-T forms counted as post-introduction documentation where available. The review described documentation, not adherence to the 4-hour target or all bedside care.

Reviewers were clinicians familiar with delirium care at their site and used locally available records. German baseline data came from delirium-team records rather than the full ward record; an absent item therefore meant it was not documented by that team, not that ward staff had not completed it. Reviewers knew the study phase, and we did not test inter-rater reliability.

### Data analysis

We summarized survey items as frequencies and percentages, using available responses as denominators with no imputation; positive ratings combined the two highest categories. We grouped free text pragmatically into recurring categories to guide revision rather than undertaking formal qualitative analysis. We summarized age as mean (standard deviation), admission-to-index time as median, and categorical variables as counts and percentages. Care-action counts and proportions were reported by site and phase, excluding missing or not-applicable entries. We did not pool effects or perform hypothesis tests because samples were small and non-random and sites differed in ward mix and record sources.

### Ethics and information governance

At Stanford, the institutional review board determined that the project did not constitute human subjects research and that no further IRB review was required (protocol 79545, "What to do about a positive delirium screen"). At Frankfurt, the institutional review board of the state of Hesse approved the use of retrospectively collected data (2026-4463-evBO). The Edinburgh component was an approved service evaluation project under local quality improvement governance. Survey responses were anonymous.

Sites removed direct patient identifiers, assigned study codes, and collated only de-identified data.

## Results

### Staff perceptions of the ACE-T tool

The structured survey was completed by 53 multidisciplinary staff (26 nurses, 19 physicians and 8 advanced practice providers). Respondents rated the ACE-T favorably overall: 75% (39/52) rated it as easy or very easy to use, 77% (41/53) rated its instructions and steps as clear or very clear, 70% (37/53) rated it as very or extremely helpful for nursing assessment and management, and 71% (37/52) rated it as very or extremely helpful for improving their current approach to delirium care.

Nevertheless, 40% (21/52) identified a section that was unclear or difficult, and 22 respondents provided free-text suggestions. Recurring suggestions were to distinguish nursing from medical actions more clearly, especially within Acute Triggers; simplify that section for a busy workflow; integrate ACE-T with existing documentation to avoid duplication; expand person-centered strategies within Patient Experience; and phrase items as questions or instructions. Specific areas of uncertainty included medication review (5/22, 23%), diagnostic work-up and laboratory-ordering boundaries (5/22, 23%), Acute Triggers (4/22, 18%), layout (4/22, 18%), and NEWS2 or QDAT (3/22, 14%). These findings informed the revisions described in the Methods.

### Pilot records and sampling context

Sixty records were reviewed (30 before and 30 after introduction; 10 per site in each phase). Mean age was similar (pre-introduction 86.2 years [SD 6.7; n=29 with age recorded] and post-introduction 85.9 years [SD 6.7; n=30]), whereas the proportion of male patients differed (63% and 43%, respectively).

Median time from admission to the index event was 1 day in both phases. Ward mix changed substantially: 70% of pre-introduction records came from general medical wards and 3% from geriatric wards, compared with 23% and 67%, respectively, after introduction. At the UK and US sites, all post-introduction records came from geriatric units. Findings are therefore reported as site-level descriptive counts rather than pooled before-and-after comparisons.

### Documentation patterns and measurement learning

The most consistent site-level pattern concerned documentation of delirium or suspected delirium (Table 3). This was present in 1/10 pre- and 10/10 post-introduction records in Frankfurt, 0/10 and 10/10 in Edinburgh, and 3/10 and 9/10 in Stanford. Communication of screening or assessment findings to the medical team was recorded in 4/10 pre- and 9/9 post-introduction applicable records in Frankfurt, 2/4 and 10/10 applicable records in Edinburgh, and 0/5 and 3/10 applicable records in Stanford. Patient or family communication and leaflet provision were more variable.

**Table 3.**
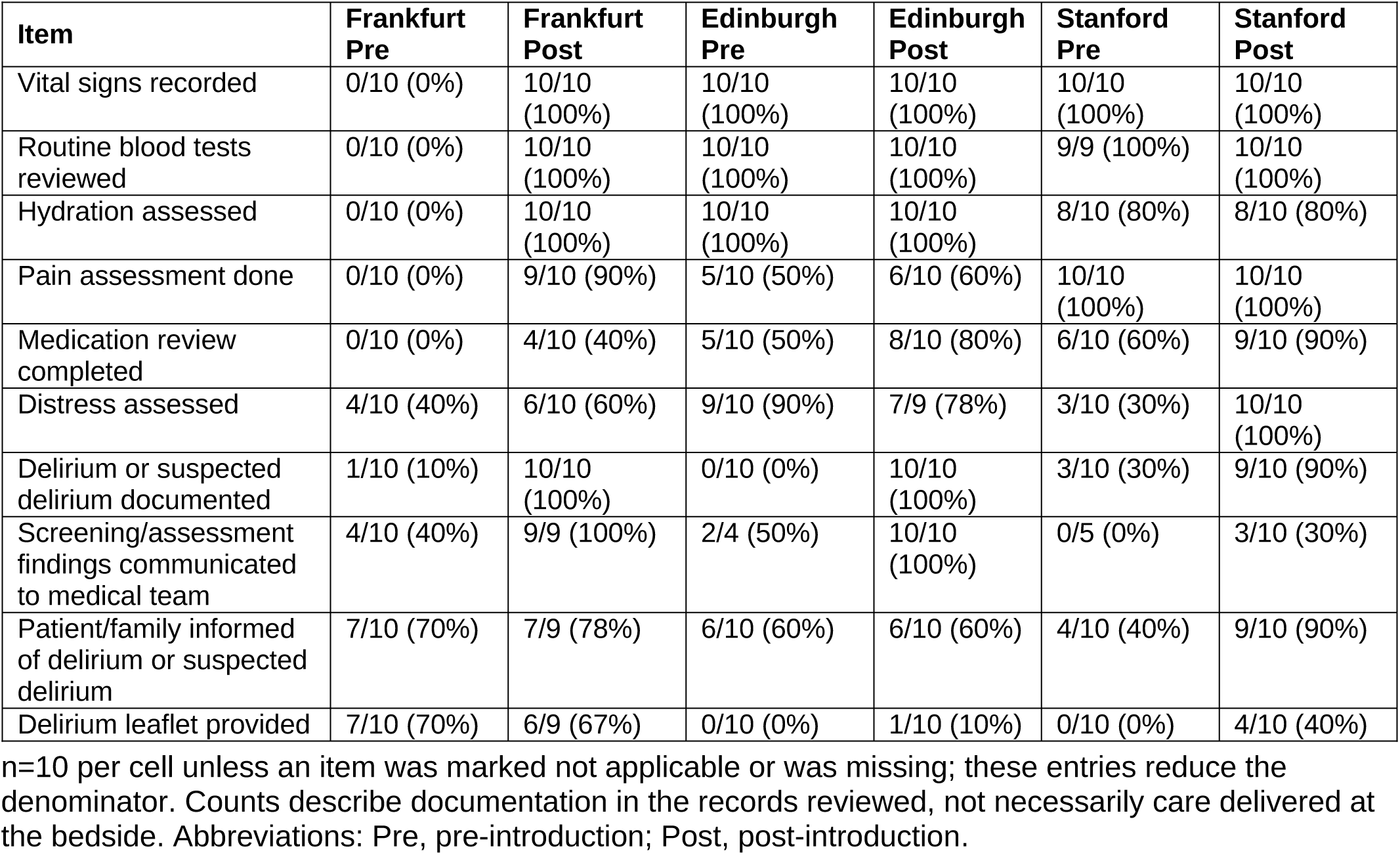
Site-stratified pre- vs post-introduction documentation for selected ACE-T care actions.

Routine assessments for potentially reversible causes and contributing factors varied across sites, with no clear overall pattern. Documentation of vital signs and blood test review was already high in Edinburgh and Stanford, and remained high. In Frankfurt, several routine items were documented in 0/10 baseline delirium-team records and 10/10 post-introduction records. Because baseline abstraction used delirium-team rather than comprehensive ward records, this difference is likely to reflect, at least in part, the source of documentation rather than absence of routine ward assessment. These values were treated as measurement learning, not evidence of a pooled improvement in care.

Documentation of distress was recorded in 3/10 pre- and 10/10 post-introduction records in Stanford, 4/10 and 6/10 in Frankfurt, and 9/10 and 7/9 in Edinburgh.

## Discussion

The ACE-T addresses a practical gap between detecting delirium and initiating a coordinated response. In this pilot, staff from several disciplines generally found the tool clear, usable and helpful, and their feedback informed changes to scope, workflow and person-centered content. Across all three sites, the record review found that delirium or suspected delirium and communication with the medical team were documented more often after introduction.

The same delirium-specific documentation pattern was seen across the three systems. Detection alone does not ensure that delirium is documented or escalated, and previous studies describe the same post-screen gap.^20,29,31-34^ The ACE-T offers a structured way to make the expected initial response explicit and more visible within the wider multidisciplinary pathway. Testing at three sites showed the need for local adaptation, and a shared framework may support a more consistent and coordinated response.

The ACE-T is an initial-response prompt focused on key nursing elements of delirium treatment. It is not a diagnostic instrument or complete treatment pathway. It prompts nurses to consider appropriate comfort, observation, assessment and person-centered measures; make possible triggers visible; document suspected delirium; communicate with the team and escalate concerns according to local policy. This is consistent with guidance recommending rapid assessment, management of acute triggers and mitigation of other contributors (such as sensory impairments), supportive care, communication and multidisciplinary management.^2,3,10-13^ Diagnosis, prescribing and definitive treatment remain multidisciplinary and scope-dependent. The 4-hour interval was a pragmatic completion target, not a known safety threshold. Urgent concerns require immediate action, and team members should work in parallel, using clinical judgement depending on the clinical situation. In Edinburgh, investigators considered the tool’s explicit nursing focus and simple, stepwise format important for local engagement and implementation.

The staged development was a strength. Feedback informed revision before pilot use, and testing in three systems identified contextual differences in professional scope, terminology, workflow and documentation that a single-site study might have missed. Current guidance on complex interventions similarly emphasizes context, stakeholder perspectives and iterative refinement.^35^

This preliminary study has limitations. Completed ACE-T forms counted as post-introduction documentation, so the study could not distinguish changes in bedside care from changes in recording. The 48-hour window did not test completion within 4 hours. German baseline data came from delirium-team records, while ward mix changed between phases in the UK and USA. Eligible-case denominators, reach, uptake and fidelity were not measured consistently; confounding and contextual differences limit direct comparisons and generalizability.

Other limitations include small convenience samples, non-random case selection, heterogeneous triggers, unblinded review, no inter-rater reliability testing and possible concurrent influences. The pre-use questionnaire was unvalidated, 26 of 53 respondents were nurses, and observed usability was not assessed. Factors supporting local engagement were not formally evaluated. Patients and family carers were not involved in initial development. The communication item did not establish the recipient, content or quality of information. Safety outcomes, unintended consequences, repeat use and staff burden were not assessed. ACE-T also depends on reliable upstream detection, which remains variable in routine practice.^8,18^

A prospective feasibility study should co-design the implementation of the tool with bedside nurses, patients and carers; examine how the ACE-T integrates with the work of other disciplines; formally assess content validity and usability; and recruit consecutive eligible cases. Measures should distinguish implementation from clinical effects and include reach, adoption, 4-hour completion, fidelity, staff burden, acceptability, patient and carer communication, and unintended consequences.^36^ Electronic integration should be tested, including whether it reduces duplicate recording without adding alert or documentation burden. Establishing feasibility and measurement methods would provide a basis for evaluating clinical effectiveness.

The ACE-T is a promising, practical tool for the initial nursing response to delirium. Its stepwise format offers a straightforward starting point for teams seeking to make these first steps clearer and more consistent in routine care. Future work will help assess its value in clinical practice.

## Supporting information

Supplementary Material: staff questionnaire and blank baseline data extraction form

## Acknowledgements

We thank colleagues who participated in the various stages of the project, including the survey and implementation elements.

## Author contributions

Conceptualization: AM, MH; Methodology: AM, MH, AC, JT; Investigation: AM, JT, AC, MH, US, ES, NSS; Formal analysis: AC; Writing - original draft: AC; Writing - review & editing: all authors. All authors reviewed and approved the final manuscript.

## Ethics statement

At Stanford, the institutional review board determined that the project did not constitute human subjects research and that no further IRB review was required (protocol 79545, "What to do about a positive delirium screen"). At Frankfurt, the institutional review board of the state of Hesse approved the use of retrospectively collected data (2026-4463-evBO). The Edinburgh component was an approved service evaluation project under local quality improvement governance. Survey participation was voluntary, and return of the questionnaire indicated consent. Survey responses were anonymous. Sites removed direct patient identifiers, assigned study codes, and collated only de-identified data.

## Data availability

De-identified aggregate data are available from the corresponding author on reasonable request, subject to local governance requirements.

