## Supplementary Material: staff questionnaire and blank baseline data extraction form for "Development and pilot evaluation of ACE-T, a nurse-focused delirium initial-response tool, across three health systems"

### ACE-T-N nursing delirium management tool: quick survey

The ACE-T-N is a provisional tool designed to guide the first steps following a positive score on a delirium detection tool (such as the 4AT or the CAM). ACE-T-N stands for Acute Triggers / patient Experience / Treatments + N (Nursing). This tool has been designed specifically to be used by nurses. Actions suggested by the ACE-T-N would inform the team approach to managing the patient's delirium, including actions by medical staff.

We would like to learn about your opinion on this tool by means of this brief survey.

Many thanks for your time.

---

**Please circle your professional discipline:**

Nurse Physician Physician assistant/associate Nurse practitioner

**Please circle how many years of experience you have caring for patients with delirium:**

0-2 years 2-5 years 5-10 years >10 years

**1. On a scale of 1 to 5 (1 = very difficult, 5 = very easy), how easy is it to understand and use the ACE-T-N?**

☐ 1 ☐ 2 ☐ 3 ☐ 4 ☐ 5

**2. How clear are the instructions and steps outlined in the ACE-T-N?**

☐ Very unclear ☐ Unclear ☐ Neutral ☐ Clear ☐ Very clear

**3. How helpful do you think the ACE-T-N would be in guiding the initial nursing assessment and management of delirium?**

☐ Not helpful ☐ Slightly helpful ☐ Moderately helpful ☐ Very helpful ☐ Extremely helpful

**4. Do you think that the ACE-T-N would help in improving the current approach to managing delirium?**

☐ Not helpful ☐ Slightly helpful ☐ Moderately helpful ☐ Very helpful ☐ Extremely helpful

**5. Are there any sections of the ACE-T-N that you find unclear or difficult to use? If yes, please specify:**

☐ No ☐ Yes (please specify):

**6. Are there any changes or additions that you would suggest to improve the ACE-T-N, or do you have any comments?**

### Delirium Quality Improvement Baseline Assessment Form

This form is designed for **retrospective hospital record review** to assess how well **ACE-T-N** (Acute Triggers, Patient Experience, Treatments, Nursing) actions have been followed within **48 hours** after a delirium diagnosis. The goal is to establish a baseline adherence level before initiating quality improvement interventions.

#### 1. General Patient Information

- Patient ID (anonymized code)
- Date of Admission: YYYY-MM-DD
- Date of Delirium Diagnosis: YYYY-MM-DD
- Age: ___ years
- Sex: ☐ Male ☐ Female ☐ Other
- Ward Type: ☐ Orthopedic ☐ Medicine of Elderly/Geriatrics ☐ General Medical ☐ Other (specify)
- Diagnosis Tool Used: ☐ 4AT ☐ CAM ☐ Other (specify)
- Delirium Subtype: ☐ Hypoactive ☐ Hyperactive ☐ Mixed ☐ Not recorded
- Primary Suspected Cause(s): ☐ Infection ☐ Surgery ☐ Medication ☐ Constipation ☐ Dehydration ☐ Hypoxia ☐ Other (specify)________________

#### 2. Acute Triggers Assessment

- Vital signs recorded? ☐ Yes ☐ No
- Blood glucose checked? ☐ Yes ☐ No
- Urinary retention assessed? ☐ Yes ☐ No
- Constipation assessed? ☐ Yes ☐ No
- Hydration status assessed? ☐ Yes ☐ No
- Nutrition status assessed? ☐ Yes ☐ No

- Recent blood tests available & reviewed? ☐ Yes ☐ No
- Any infection workup done? ☐ Yes ☐ No | Specify (urinalysis, chest X-ray, etc.)
- Pain assessment done? ☐ Yes ☐ No
- Medication review completed? ☐ Yes ☐ No | Deliriogenic meds identified? ☐ Yes ☐ No

#### 3. Patient Experience Assessment

- Distress assessed? ☐ Yes ☐ No | If yes, was reassurance provided? ☐ Yes ☐ No
- Patient's environment considered? ☐ Yes ☐ No

  If needed, was a single room arranged? ☐ Yes ☐ No
- Communication of delirium diagnosis to family? ☐ Yes ☐ No
- Delirium leaflet provided? ☐ Yes ☐ No

#### 4. Nursing Documentation & Communication

- Delirium diagnosis clearly documented? ☐ Yes ☐ No
- 4AT / CAM Score Recorded? ☐ Yes ☐ No
- Findings communicated to medical team? ☐ Yes ☐ No
- Findings communicated to patient and/or family (if appropriate)? ☐ Yes ☐ No

#### 5. Additional Observations & Reviewer Comments

- (Use this section to record any additional notes, observations, or discrepancies in care observed within 48 hours post-diagnosis.)
